# From district to community: expanded eligibility, not wider geography, drives schistosomiasis programme costs under revised WHO guidance in Ethiopia and Zimbabwe

**DOI:** 10.64898/2026.04.07.26350372

**Authors:** Alexandra Carlin, Catherine Fantaguzzi, Fikre Seife, Gemechu T. Leta, Isaac Phiri, Neerav Dhanani, Nicholas Midzi, Fiona M Fleming

## Abstract

**Background:** Schistosomiasis remains a major public health challenge in sub-Saharan Africa. Recent World Health Organization (WHO) guidance calls for community-wide treatment and fine-scale data to optimise preventive chemotherapy (PC) strategies, yet the practical implications for resource allocation by health ministries are unclear.

**Methods:** We analysed epidemiological and cost data from Ethiopia and Zimbabwe to compare survey designs and five implementation scenarios. Scenarios varied by data source, administrative unit of implementation, and WHO guidance on PC strategies. Outcomes were target population, praziquantel requirements, and delivery costs.

**Results:** The geostatistical approach reduced sample size by up to 90% and survey costs by ≥72% compared with a design-based approach, while increasing spatial coverage. Applying updated WHO guidance expanded eligibility to pre-school-aged children and adults, and in one scenario increased treatment eligibility by 72% in Ethiopia and 262% in Zimbabwe. Correspondingly, praziquantel requirements and delivery costs were driven primarily by expanded age eligibility rather than geographic coverage.

**Conclusions:** The geostatistical approach provides substantial efficiency gains for impact assessments, enabling cost-efficient, granular targeting. However, implementing 2022 WHO guidance was the dominant driver of increases in programme scope and resource needs, underscoring the importance of accurate fine-scale data to guide efficient planning and budgeting toward elimination goals.

**Author summary:** Schistosomiasis control programmes are required to use finer-scale data and updated World Health Organization (WHO) guidance to decide where and how often to deliver praziquantel. We analysed national schistosomiasis data and programme costs from Ethiopia and Zimbabwe to compare different approaches to impact assessment surveys and to estimate how the population requiring preventive chemotherapy changes under alternative decision rules. We found that a geostatistical approach can reduce the number of people that need to be sampled and the cost of surveys while providing more detailed information for planning at sub-implementation unit (sub-IU) level. However, when we applied the 2022 WHO schistosomiasis guidance, expanded eligibility (including adults and pre-school-age children and a lower threshold for community-wide treatment) substantially increased the number of people needing treatment. In our scenarios, expanded eligibility drove much larger increases in praziquantel requirements and delivery costs than changes in geographic coverage. Our findings help health ministries to anticipate the operational and budget implications of updated guidance and highlight why accurate fine-scale data are essential for equitable and realistic planning toward elimination.

## Introduction

Schistosomiasis continues to pose a major public health problem in the African region [1]. Caused by *Schistosoma* blood flukes, schistosomiasis contributes an estimated 1.8 million disability-adjusted life-years (DALYs) annually through anaemia, chronic inflammation, and intestinal, hepatosplenic, and urogenital disease [2]. Control has focused on mass preventive chemotherapy (PC) with praziquantel (PZQ), with programmes expanding over the past 20 years in endemic areas [1,3,4]. Recent analyses have indicated widespread prevalence declines across sub-Saharan Africa between 2000 and 2019, likely attributable to PC [4]. To consolidate these gains, the World Health Organization’s (WHO) 2021-2030 Neglected Tropical Diseases (NTD) Roadmap aims to eliminate schistosomiasis as a public health problem (EPHP) by 2030 [5].

Schistosomiasis-specific WHO guidelines require programmes to inform and adapt PC strategies with epidemiological prevalence data through baseline mapping prior to interventions and periodic impact assessments thereafter [6–8]. To account for the observed heterogeneity in schistosomiasis distribution, the NTD Roadmap also recommends a shift from district-to community-level PC implementation which requires infection risk to be delineated at much finer spatial scales [9–12]. For example, prevalence of infection— previously determined at an implementation unit (IU), such as a district or health district from baseline— would now be used to make decisions at a sub-IU, such as ward, kebele, or community. Consequently, finer-scale epidemiological data will enable better targeting of schistosomiasis control interventions such as PC, minimising both under- and over-treatment [11,13]. Additionally, from 2022 all endemic countries applying for donated PZQ through the WHO partnership with Merck KGaA’s Schistosomiasis Elimination Program are requested to do so based on sub-IU prevalence and to implement and report at this level [14].

Providing fine-scale estimation for a focal disease such as schistosomiasis is challenging without substantial data collection. Health ministry NTD programmes have tested multiple design-based survey approaches for impact assessments to describe epidemiology at the sub-IU, or below [9,12,13,15–19]. Ethiopia and Zimbabwe, with estimated baseline national schistosomiasis prevalence of 4.0% and 22.7%, respectively, decided to conduct their national impact assessments using a geostatistical approach [20,21]. The geostatistical approach for both design and analysis was selected by the health ministries due to its sampling efficiency, whilst ensuring statistical rigour, as compared to design-based cluster surveys [11,22].

The geostatistical approach to impact assessments provided the Ethiopian and Zimbabwean health ministries with precise prevalence classifications at the sub-IU (kebele and ward, respectively). Impact assessment data were then used to support decisions on which populations to target with priority interventions, including PC. At the time of conducting the data collection surveys in Ethiopia and Zimbabwe (2019-24), existing WHO algorithms were in place to support post-impact assessment decision-making for PC frequency and target populations for schistosomiasis [8]. During the period of impact assessment analysis (2022-23), new WHO guidelines were released which updated the PC decision-making thresholds and algorithms [6]. The new guidelines included widening the target population from school-aged children (SAC) and at-risk adults in high prevalence areas to whole communities from two years of age and lowering the prevalence threshold for this community-wide treatment to 10% prevalence, from 50%, for annual PC with PZQ.

With these simultaneous changes to decision-making rules and requirements for more precise epidemiological data, we hypothesised that (1) a geostatistical approach would reduce sample size and survey cost, relative to a design-based survey, while improving spatial targeting; (2) moving decision-making from the IU to the sub-IU would allow PC to be reduced or increased in more focal areas, reducing the geographic extent of annual PC; and (3) the 2022 WHO guidelines would show an expansion in the total population eligible for PC and hence PZQ requirements and delivery cost, offsetting any geographic shrinkage of need. This study aimed to use epidemiological and cost evidence from Ethiopia and Zimbabwe, to demonstrate efficiency gains in survey design and to determine differences in target populations and costs when using (i) more precise epidemiological data from impact assessments, (ii) historical and updated WHO guidelines, and (iii) IU versus sub-IU as a unit for informing intervention decisions. These results increase the evidence in understanding optimal survey designs and resource allocation and are the first to demonstrate the impact of policy changes in the implementation of national control and elimination programmes for schistosomiasis in sub-Saharan Africa.

## Methods

Historically, Ethiopia and Zimbabwe health ministry programmes have implemented at the IU-level, which is woreda and district, respectively and shifted to sub-IU schistosomiasis PC, targeting kebele and ward based on recent prevalence data. The geostatistical models underpinning the sub-IU prevalence classifications were developed, validated and reported as part of the respective national impact assessments [23,24]; they were not re-estimated for this study. Model specification, covariate selection and measures of predictive performance are reported in those publications. The analyses presented here take the published sub-IU prevalence classifications as fixed inputs and apply WHO decision algorithms and programme cost data to them. The geostatistical approach impact assessments determined sub-IU prevalence and were conducted in Ethiopia 2019-24 and in 2021 in Zimbabwe. All data were shared with the research team for the purposes of this study.

### Efficiency gains by survey design

Impact assessments were costed as surveys using either a design-based cluster approach, which was not conducted and is costed here as a hypothetical comparator, or the geostatistical approach that was implemented. Both survey designs are recommended by WHO for impact assessments [7].

Costs were estimated assuming several parameters. Sample sizes for the design-based survey assumed 30 SAC per site with five sites randomly selected per suspected or known endemic sub-IU. Sub-IUs not suspected or known to be endemic were not sampled and were classified as non-endemic. In the design-based survey analysis, inaccessible areas due to insecurity and political instability were excluded from the sub-IU estimates in Ethiopia. The design-based parameters were set to a 95% confidence interval, 85% power, a 10% margin of error, and an intraclass correlation coefficient of 0.10 to provide a precise estimate of sub-IU prevalence following the parameters derived by Knowles et al. for cluster surveys designed to plan schistosomiasis PC [25].

The geostatistical approach exploits the fact that prevalence at nearby locations tends to be similar: a model fitted to the sampled sites, together with environmental and demographic covariates, predicts prevalence with quantified uncertainty at unsampled locations, so that every sub-IU can be classified without sampling each one [22]. For the geostatistical survey in Ethiopia, 30 SAC per site were sampled across nationally representative sites selected using spatially regulated sampling with minimum distance of 10 km and 50% close pairs [23]. Predictions, based on multiple data sources—empirical epidemiological and environmental and demographic covariates—were then used to classify each sub-IU into WHO schistosomiasis prevalence categories using the 1%, 10% and 50% thresholds [7,23]. Applying the same classification framework, Zimbabwe leveraged extensive epidemiological data available from a 2018 survey to inform their geostatistical impact assessment, sampling 58 SAC per site [24, 26]. Sites were systematically selected with inclusion probability proportional to the estimated likelihood of exceeding 2% prevalence for *S. haematobium* in 2018 [24]. These per-site sample sizes were determined by simulation. In Ethiopia, a 2020 parameter calibration survey showed no material gain in parameter recovery beyond 30 children per school [23]; in Zimbabwe, design simulations using 2018 survey data identified 200 primary sampling units of 58 children as classifying 90% of wards correctly with least survey effort [24]. Full design specifications are reported in [23,24]. For Ethiopia, where areas could not be sampled due to political insecurity, kebele that lay greater than 20 kilometres from a sampled school site were excluded due to the increased uncertainty in correctly classifying the sub-IU. This threshold was based on the upper 95th centile of distances between survey sites and the estimated range of the spatial Gaussian process from the fitted models [23].

Efficiency gains due to survey design were then evaluated by comparing the design-based to the geostatistical approach on two key metrics (i) sample population and (ii) total survey costs. Costs by survey category are summarised in Supplementary Table S1. Cost per sub-IU was determined by dividing total costs by the number of sub-IUs with prevalence estimates.

### Efficiency gains in implementation

To determine the efficiency gains in the implementation of the Ethiopian and Zimbabwean national programmes, we tested five scenarios. The scenario decision-variables were (i) data source (baseline and the geostatistical approach impact assessment); (ii) administrative unit of implementation (IU or sub-IU); and (iii) WHO guidance on PC algorithm thresholds and strategies for schistosomiasis (Table 1). In our analysis we included treatments for pre-SAC (pre-school-aged children) and adults, however the current PZQ donation is limited for adult populations, and the new paediatric arpraziquantel is currently being piloted for optimal delivery models [27]. Thus, study outputs may not be representative of actual national target populations and PZQ requests.

**Table 1.**
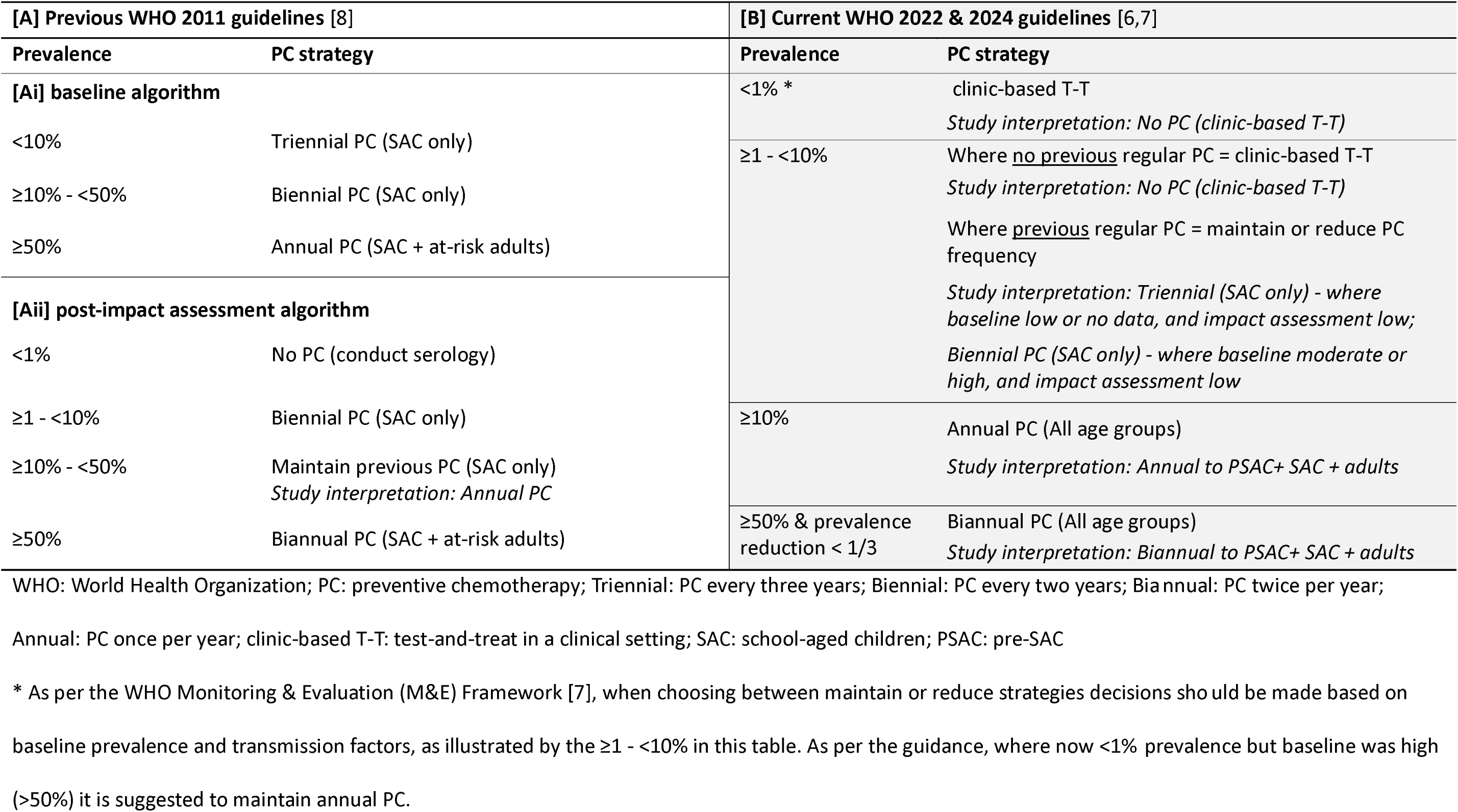
World Health Organization preventive chemotherapy algorithm thresholds and strategies for schistosomiasis, [A] 2011 [8] and [B] 2022 [6,7]

| [A] Previous WHO 2011 guidelines [8] |  | [B] Current WHO 2022 & 2024 guidelines [6,7] |  |
|---|---|---|---|
| Prevalence | PC strategy | Prevalence | PC strategy |
| [Ai] baseline algorithm |  | <1% * | clinic-based T-T<br>*Study interpretation: No PC (clinic-based T-T)* |
| <10% | Triennial PC (SAC only) | ≥1 - <10% | Where no previous regular PC = clinic-based T-T<br>*Study interpretation: No PC (clinic-based T-T)*<br>Where previous regular PC = maintain or reduce PC frequency<br>*Study interpretation: Triennial (SAC only) - where baseline low or no data, and impact assessment low;*<br>*Biennial PC (SAC only) - where baseline moderate or high, and impact assessment low* |
| ≥10% - <50% | Biennial PC (SAC only) |  |  |
| ≥50% | Annual PC (SAC + at-risk adults) |  |  |
| [Aii] post-impact assessment algorithm |  |  |  |
| <1% | No PC (conduct serology) |  |  |
| ≥1 - <10% | Biennial PC (SAC only) | ≥10% | Annual PC (All age groups)<br>*Study interpretation: Annual to PSAC+ SAC + adults* |
| ≥10% - <50% | Maintain previous PC (SAC only)<br>*Study interpretation: Annual PC* |  |  |
| ≥50% | Biannual PC (SAC + at-risk adults) | ≥50% & prevalence reduction < 1/3 | Biannual PC (All age groups)<br>*Study interpretation: Biannual to PSAC+ SAC + adults* |

WHO: World Health Organization; PC: preventive chemotherapy; Triennial: PC every three years; Biennial: PC every two years; Bia nnual: PC twice per year; Annual: PC once per year; clinic-based T-T: test-and-treat in a clinical setting; SAC: school-aged children; PSAC: pre-SAC
\* As per the WHO Monitoring & Evaluation (M&E) Framework [7], when choosing between maintain or reduce strategies decisions sho uld be made based on baseline prevalence and transmission factors, as illustrated by the ≥1 - <10% in this table. As per the guidance, where now <1% prevalence but baseline was high (>50%) it is suggested to maintain annual PC.

The scenario outcomes measured were (a) number of sub-IU, total population, and by target age-category i.e. pre-SAC, SAC and adults that would require PC, (b) number of PZQ tablets required, and (c) total treatment cost based on the delivery cost of PC and excluding drug costs.

Scenario One kept the IU constant and followed 2011 WHO PC algorithm thresholds using baseline data then using impact assessment data to support PC decisions, highlighting change in epidemiological data. Scenario Two compared “the old to new”, at IU-level, changing the epidemiological data from baseline to impact assessment and WHO PC algorithm thresholds from 2011 to 2022 guidance. Scenario Three explored the implications of changing the administration unit from IU to the sub-IU, keeping constant epidemiological data (impact assessment) and 2022 WHO PC algorithm thresholds. Scenario Four assessed the impact of updated WHO guidelines by applying the different PC algorithm thresholds and strategies to impact assessment epidemiological data at sub-IU. Scenario Five compared the impact of changing all decision-variables and reflects the current situation of health ministries, moving from PC strategies using baseline data, at the IU level, using the previous WHO guidelines to using impact assessment data, at the sub-IU level, using the current WHO guidelines. Scenarios are summarised in Table 2.

**Table 2.** Summary of five implementation scenarios with changes in (i) data source (baseline and the geostatistical approach impact assessment); (ii) administrative unit of implementation (implementation unit (IU) or sub-IU); and (iii) World Health Organization guidance on preventive chemotherapy algorithm thresholds and strategies for schistosomiasis.

|  | **a** | **b** | **c** | **d** | **e** |  |
|---|---|---|---|---|---|---|
| Data source<br>Administrative unit<br>WHO guidance (Table 1) | baseline<br>IU<br>[Ai] | impact<br>IU<br>[Aii] | impact<br>IU<br>[B] | impact<br>sub-IU<br>[Aii] | impact<br>sub-IU<br>[B] | difference |
| **Scenario one**<br>(data source) |  |  |  |  |  | b- a |
| **Scenario two**<br>(data source + guidance) |  |  |  |  |  | c -a |
| **Scenario three**<br>(administrative unit) |  |  |  |  |  | e - c |
| **Scenario four**<br>(guidance) |  |  |  |  |  | e - d |
| **Scenario five** (data source + guidance + administrative unit) |  |  |  |  |  | e - a |

WHO: World Health Organization; IU: implementation unit; sub-IU: sub-implementation unit; [Ai], [Aii] and [B]: WHO guidance (see Table 1). Shaded cells indicate the columns compared in each scenario.

All target populations and PZQ tablet quantities for each PC strategy were calculated aligned to the WHO’s Joint Request for Selected PC Medicines (JRSM) [28]. For instance: biannual PC, total target population x two; annual PC, total target population x one; biennial, total target population ÷ two; triennial, total target population ÷ three; at-risk adults, 20% of the adult population. The number of PZQ tablets were calculated using two tablets per SAC and three tablets per adult for PZQ [7]. Pre-SAC were not included in praziquantel tablet requirements, as they would be treated with paediatric arpraziquantel [27]. Population estimates were sourced from WorldPop’s top-down datasets (Constrained Individual countries 2020 100m resolution, accessed 28-11-2025) [29], as comparable sub-IU census data were not consistently available across both countries at the time of analysis.

Design-based baseline mapping surveys to determine IU prevalence were conducted in Ethiopia in 2013-15 and in Zimbabwe from 2010-11 [20,21]. The data from these surveys were obtained from the JRSM for Ethiopia and published literature for Zimbabwe. Records were checked for duplicate entries, implausible prevalence values and consistency of administrative identifiers before analysis.

### Cost data

Survey costs were estimated with survey budgets and financial expenditure reports, using an ingredients-based approach, including unit costs, quantities, or days [30]. Items were compiled into standard cost categories (e.g. staff salaries, per diems, transport and fuel, survey consumables, results dissemination workshops). Capital items were annualised based on their expected lifespan, applying a 10% wastage rate [31]. Costs assumed no existing national stockpiles of survey materials. As the design-based impact assessments were hypothetical, we used prior field experience, existing budgets, expenditure data, and published literature to assume parallel survey team size, and that survey duration changed with sample size. Survey duration was then used to estimate duration-dependent cost categories (e.g., fuel).

Treatment costs included the cost of delivering PZQ but not the drug itself due to the donation programme [32]. The average cost per delivery (CPD) was calculated as total programme annual expenditure/total annual treatments delivered. For Ethiopia, the CPD was US$0.20 and for Zimbabwe, US$0.44. Target populations were multiplied by their respective CPD to estimate total delivery costs. All costs were converted to 2024 US dollars using an exchange rate of 1 GBP = 1.22 US dollar [33].

### Key assumptions

To determine the treatment populations, decision-rules used in the WHO PC algorithm thresholds and strategies (Table 1) were used. For Table 1[B], we applied decision-rules aligned to WHO’s Community Data Analysis Tool (CDAT) version 6 [34]. Several additional interpretations were made.

For 2011 guidelines post-impact assessment (Table 1 [Aii]), where prevalence was ≥10% and <50% the PC strategy is “maintain previous frequency”. We interpreted this as annual PC based on the guidance in WHO tools (e.g., JRSM), which suggest using a continuation of the previous PC strategy (where ≥50% prevalence) or increased PC frequency (from biennial to annual) due to either an increase or no change to baseline prevalence (≥10% - <50%). For 2022 guidelines (Table 1 [B]), multiple interpretations were informed by the WHO’s CDAT PC algorithms. For instance, a clinic-based test-and-treat (T-T) approach is recommended by WHO where there was previously no regular PC; we also applied this rule to <1% prevalence areas and calculated no PC. Where prevalence ≥1% and <10% and prior regular PC, a ‘maintain or reduce PC’ strategy is recommended. For this, the CDAT generally applies triennial PC to all sub-IUs, regardless of previous strategy, with countries choosing a triennial or biennial approach during CDAT verification. We used triennial where baseline was low or no data, and impact assessment low and biennial where baseline was moderate or high, and impact assessment low, as the CDAT allows this choice at verification and biennial is the more precautionary option where baseline prevalence was moderate or high.

Sub-IU prevalence categories were assigned in the primary surveys on the basis of exceedance probability rather than a single predicted value: 1,000 realisations were simulated from the predictive distribution, and a sub-IU was assigned to a category where the probability of exceeding its lower bound was at least 30%, a threshold chosen to balance the risk of under-treatment against the cost of over-treatment [23,24].

The geostatistical approach impact assessment outputs are provided as sub-IU prevalence categories (e.g. <1%, ≥1% to <10%, etc). When aggregating IU prevalence from these results for these analyses, an IU was classified as ≥10% prevalence if more than 10% of its sub-IUs exceeded 10%. A low threshold was applied deliberately, so that an IU containing a minority of higher-prevalence sub-IUs was not classified below the treatment threshold, consistent with the precautionary basis of the exceedance probability threshold applied in the primary surveys as advised by health ministries [23,24].

### Data analysis

No statistical estimation was undertaken in this study. Scenario outputs are deterministic functions of the published sub-IU prevalence classifications, the WHO decision algorithms (Table 1), population denominators and unit costs, and no sampling distribution is associated with them; results are therefore presented as point estimates. Costing calculations were carried out in Microsoft Excel and scenario calculations in R version 4.2 using the dplyr and tidyr packages. Model fitting, validation and predictive uncertainty for the underlying geostatistical surfaces are reported in the primary survey publications [23,24]. Analysis code and scenario input files are available on request.

### Sensitivity analysis

To assess the robustness of the survey cost comparison, we conducted a one-way sensitivity analysis. Each cost category (Supplementary Table S1) was varied by ±25%, applied to both survey types simultaneously as both were costed from the same unit costs. Technical support was additionally varied by ±50%, as the input most specific to the geostatistical approach. We also varied the wastage rate applied to capital items from 0% to 20%. The outcome was the percentage difference in total survey cost between survey types (Supplementary Table S2). Treatment costs were not varied, as target population, praziquantel requirements and delivery costs are each a fixed multiple of the preceding quantity; a proportional change in the cost per delivery or in population estimates therefore alters all scenarios equally and leaves the comparisons between them unchanged.

### Ethical approval

All analysis for this study was conducted on de-identified secondary data and exempt from ethical review. The health ministries of both countries approved the use of the anonymised data. Ethical approval for the original surveys was obtained from Ethiopian Public Health Institute Institutional Review Board (EPHI-IRB-368-2021) and Medical Research Council of Zimbabwe (MRCZ/A/2810).

## Results

### Efficiency Gains by Survey Design

Marked differences were observed between the design-based and geostatistical approach (Table 3). In Ethiopia, the design-based survey estimated a sample population of 2.11 million individuals, whereas the geostatistical approach required 201,990 individuals, a 90.4% reduction. The geostatistical approach generated prevalence estimates in Ethiopia for 19,475 sub-IUs compared to 10,712 sub-IUs using design-based surveys, an 81.8% increase. In Zimbabwe, the sample population fell by 90.1%, and both approaches provided estimates for all 1,961 wards.

**Table 3.**
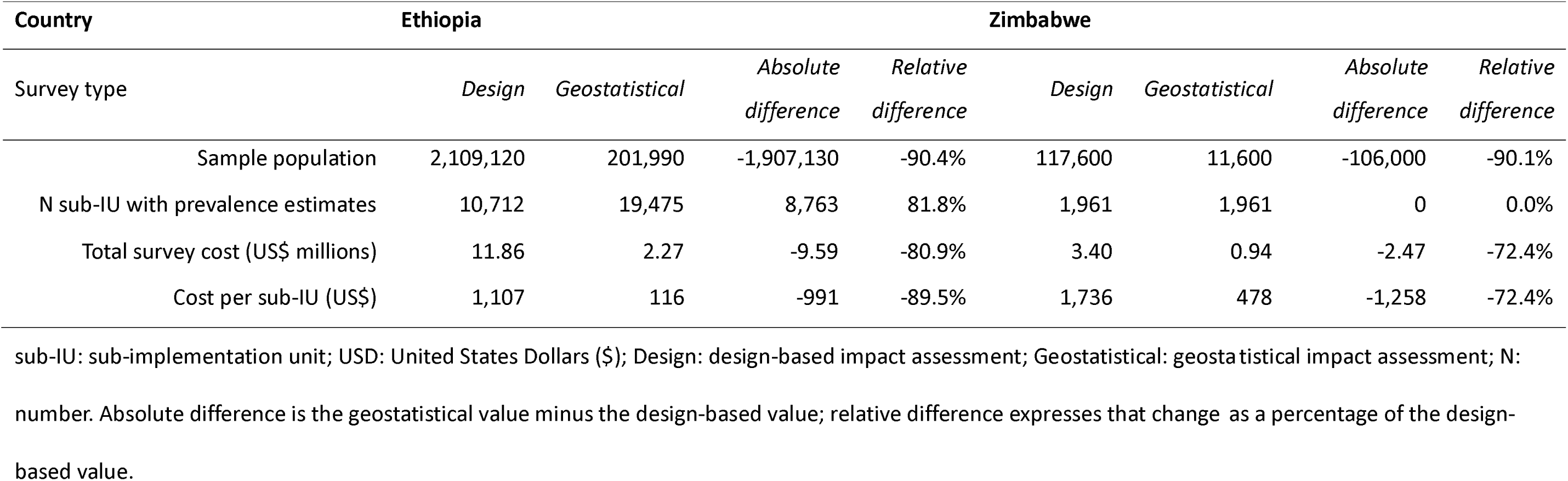
Differences in sample population, number of sub-IU with prevalence estimates, total surveys costs (USD) and cost of survey per sub-IU (USD), by survey type in Ethiopia and Zimbabwe to determine sub-IU prevalence.

| **Country** | **Ethiopia** |  |  |  | **Zimbabwe** |  |  |  |
|---|---|---|---|---|---|---|---|---|
| Survey type | *Design* | *Geostatistical* | *Absolute difference* | *Relative difference* | *Design* | *Geostatistical* | *Absolute difference* | *Relative difference* |
| Sample population | 2,109,120 | 201,990 | -1,907,130 | -90.4% | 117,600 | 11,600 | -106,000 | -90.1% |
| N sub-IU with prevalence estimates | 10,712 | 19,475 | 8,763 | 81.8% | 1,961 | 1,961 | 0 | 0.0% |
| Total survey cost (US\$ millions) | 11.86 | 2.27 | -9.59 | -80.9% | 3.40 | 0.94 | -2.47 | -72.4% |
| Cost per sub-IU (US\$) | 1,107 | 116 | -991 | -89.5% | 1,736 | 478 | -1,258 | -72.4% |

sub-IU: sub-implementation unit; USD: United States Dollars ($); Design: design-based impact assessment; Geostatistical: geostatistical impact assessment; N: number. Absolute difference is the geostatistical value minus the design-based value; relative difference expresses that change as a percentage of the design-based value.

In both countries, the geostatistical approach generated substantial cost savings, reducing survey costs by 80.9% in Ethiopia and 72.4% in Zimbabwe, and lowering overall survey costs by approximately US$12.1 million and reducing cost per sub-IU. Notably, however, cost allocation for technical support was higher when using geostatistics, reflecting the additional analytical and modelling expertise required (Supplementary Table S1).

The difference in survey costs between survey types was robust to the costing assumptions (Supplementary Table S2). Varying all cost categories simultaneously by ±25% left the difference unchanged. Varied individually by ±25%, no single category changed it by more than 1.6 percentage points and varying technical support by ±50% changed it by 2.1 percentage points. Across all variations, the reduction in survey cost using the geostatistical approach remained between 78.8% and 83.0% in Ethiopia and between 70.4% and 74.6% in Zimbabwe.

### Efficiency Gains by Implementation

#### Scenario One. Effect of a change in epidemiological data

Scenario one assessed the effect of changing epidemiological data under 2011 guidelines, comparing baseline and impact assessment data at the IU-level (Table 2). This scenario showed substantial differences in the estimated populations requiring PC for schistosomiasis in both Ethiopia and Zimbabwe, more than halving the number of sub-IUs considered endemic for schistosomiasis in Ethiopia from 7,902 to 3,561 (-54.9%) and increasing by 9.3% in Zimbabwe (1,569 to 1,715) (Table 4). The total population requiring PC correspondingly decreased in Ethiopia (-50.9%) and increased in Zimbabwe (30.9%).

**Table 4.**
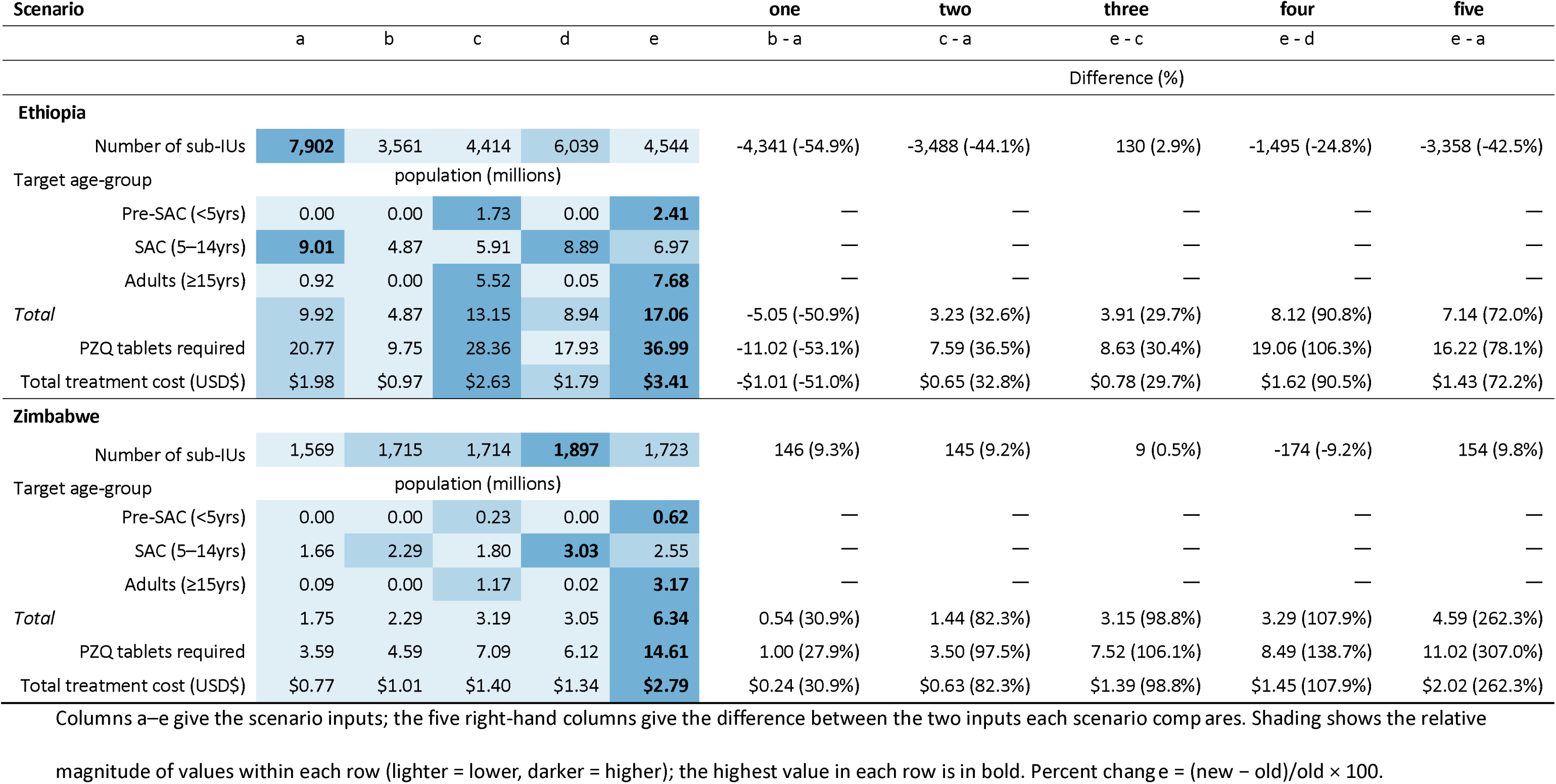
Number of sub-implementation units and population by age groups requiring preventive chemotherapy for schistosomiasis, praziquantel tablets required and total treatment cost in Ethiopia and Zimbabwe, under five scenarios. Differences show absolute change and percent change; percent change = (new-old)/old × 100; Differences for the individual age groups are reported in Supplementary Table S3.

| **Scenario** |  |  |  |  |  | **one** | **two** | **three** | **four** | **five** |
|---|---|---|---|---|---|---|---|---|---|---|
|  | a | b | c | d | e | b - a | c - a | e - c | e - d | e - a |
|  |  |  |  |  |  | Difference (%) |  |  |  |  |
| **Ethiopia** |  |  |  |  |  |  |  |  |  |  |
| Number of sub-IUs | **7,902** | 3,561 | 4,414 | 6,039 | 4,544 | -4,341 (-54.9%) | -3,488 (-44.1%) | 130 (2.9%) | -1,495 (-24.8%) | -3,358 (-42.5%) |
| Target age-group | population (millions) |  |  |  |  |  |  |  |  |  |
| Pre-SAC (<5yrs) | 0.00 | 0.00 | 1.73 | 0.00 | **2.41** | — | — | — | — | — |
| SAC (5–14yrs) | **9.01** | 4.87 | 5.91 | 8.89 | 6.97 | — | — | — | — | — |
| Adults (≥15yrs) | 0.92 | 0.00 | 5.52 | 0.05 | **7.68** | — | — | — | — | — |
| *Total* | 9.92 | 4.87 | 13.15 | 8.94 | **17.06** | -5.05 (-50.9%) | 3.23 (32.6%) | 3.91 (29.7%) | 8.12 (90.8%) | 7.14 (72.0%) |
| PZQ tablets required | 20.77 | 9.75 | 28.36 | 17.93 | **36.99** | -11.02 (-53.1%) | 7.59 (36.5%) | 8.63 (30.4%) | 19.06 (106.3%) | 16.22 (78.1%) |
| Total treatment cost (USD\$) | \$1.98 | \$0.97 | \$2.63 | \$1.79 | **\$3.41** | -\$1.01 (-51.0%) | \$0.65 (32.8%) | \$0.78 (29.7%) | \$1.62 (90.5%) | \$1.43 (72.2%) |
| **Zimbabwe** |  |  |  |  |  |  |  |  |  |  |
| Number of sub-IUs | 1,569 | 1,715 | 1,714 | **1,897** | 1,723 | 146 (9.3%) | 145 (9.2%) | 9 (0.5%) | -174 (-9.2%) | 154 (9.8%) |
| Target age-group | population (millions) |  |  |  |  |  |  |  |  |  |
| Pre-SAC (<5yrs) | 0.00 | 0.00 | 0.23 | 0.00 | **0.62** | — | — | — | — | — |
| SAC (5–14yrs) | 1.66 | 2.29 | 1.80 | **3.03** | 2.55 | — | — | — | — | — |
| Adults (≥15yrs) | 0.09 | 0.00 | 1.17 | 0.02 | **3.17** | — | — | — | — | — |
| *Total* | 1.75 | 2.29 | 3.19 | 3.05 | **6.34** | 0.54 (30.9%) | 1.44 (82.3%) | 3.15 (98.8%) | 3.29 (107.9%) | 4.59 (262.3%) |
| PZQ tablets required | 3.59 | 4.59 | 7.09 | 6.12 | **14.61** | 1.00 (27.9%) | 3.50 (97.5%) | 7.52 (106.1%) | 8.49 (138.7%) | 11.02 (307.0%) |
| Total treatment cost (USD\$) | \$0.77 | \$1.01 | \$1.40 | \$1.34 | **\$2.79** | \$0.24 (30.9%) | \$0.63 (82.3%) | \$1.39 (98.8%) | \$1.45 (107.9%) | \$2.02 (262.3%) |

Columns a–e give the scenario inputs; the five right-hand columns give the difference between the two inputs each scenario comp ares. Shading shows the relative magnitude of values within each row (lighter = lower, darker = higher); the highest value in each row is in bold. Percent chang e = (new − old)/old × 100.

WHO: World Health Organization; baseline: design-based baseline survey; impact: geostatistical approach impact assessment; IU: implementation unit; sub-IU: sub-implementation unit; [Ai], [Aii] and [B]; WHO guidance (see Table 1); SAC: school-aged children; PZQ: praziquantel; USD: United States Dollars ($) in millions

In Ethiopia, the change observed is driven by a decrease in prevalence ≥1% from baseline (Table 5), whereas the increase in Zimbabwe is due to a 0.63 million (38.0%) increase in SAC population requiring PC due to increasing PC frequency from triennial to biennial for sub-IU <10% prevalence (Table 1). No IUs were classified as ≥50% prevalence in either country, removing PC requirements for at-risk adults (Table 5).

**Table 5.** Number of sub-implementation units requiring preventive chemotherapy for schistosomiasis, by prevalence category in Ethiopia and Zimbabwe. Differences across the five scenarios are shown as absolute change and percent change, where percent change = (new-old)/old × 100; NA where old=0.

| Scenario |  |  |  |  |  | one | two | three | four | five |
|---|---|---|---|---|---|---|---|---|---|---|
| Decision-variables | a | b | c | d | e | b - a | c - a | e - c | e - d | e - a |
| Prevalence category | Number of sub-IUs |  |  |  |  | Difference (%) |  |  |  |  |
| **Ethiopia** |  |  |  |  |  |  |  |  |  |  |
| Non-endemic / <1% | 0 | 0 | 0 | 0 | 285 | 0 (NA) | 0 (NA) | 285 (NA) | 285 (NA) | 285 (NA) |
| ≥1% - <10% | 3,807 | 3,316 | 4,169 | 4,812 | 3,032 | -491 (-12.9%) | 362 (9.5%) | -1,137 (-27.3%) | -1,780 (-37.0%) | -775 (-20.4%) |
| ≥10% - <50% | 2,819 | 245 | 245 | 1,179 | 1,179 | -2,574 (-91.3%) | -2,574 (-91.3%) | 934 (381.2%) | 0 (0.0%) | -1,640 (-58.2%) |
| ≥50% | 1,276 | 0 | 0 | 48 | 48 | -1,276 (-100.0%) | -1,276 (-100.0%) | 48 (NA) | 0 (0.0%) | -1,228 (-96.2%) |
| **Zimbabwe** |  |  |  |  |  |  |  |  |  |  |
| Non-endemic / <1% | 0 | 0 | 0 | 0 | 0 | 0 (NA) | 0 (NA) | 0 (NA) | 0 (NA) | 0 (NA) |
| ≥1% - <10% | 362 | 1,503 | 1,502 | 1,183 | 1,009 | 1,141 (315.2%) | 1,140 (314.9%) | -493 (-32.8%) | -174 (-14.7%) | 647 (178.7%) |
| ≥10% - <50% | 1,058 | 212 | 212 | 698 | 698 | -846 (-80.0%) | -846 (-80.0%) | 486 (229.3%) | 0 (0.0%) | -360 (-34.0%) |
| ≥50% | 149 | 0 | 0 | 16 | 16 | -149 (-100.0%) | -149 (-100.0%) | 16 (NA) | 0 (0.0%) | -133 (-89.3%) |

WHO: World Health Organization; baseline: design-based baseline survey; impact: geostatistical approach impact assessment; IU: implementation unit; sub-IU: sub-implementation unit; [Ai], [Aii] and [B]; WHO guidance (see Table 1). Shading indicates the direction of change: blue, a decrease; orange, an increase; grey, no change.

#### Scenario Two. Effect of a change in epidemiological data plus WHO recommendations

Scenario two assessed the combined effect of updated epidemiological data and WHO guidelines while maintaining implementation at IU level (Table 2). The number of sub-IUs requiring PC resulted in a reduction of 44.1% in Ethiopia (7,902 to 4,414) and a 9.2% increase in Zimbabwe (1,569 to 1,714) (Table 4). The geostatistical approach increased spatial coverage and detection of endemic areas in Ethiopia (Table 3). Consequently, this resulted in a reduced number of sub-IUs requiring PC through the reclassification of sub-IU from <10% triennial to <1% and no PC (Tables 1, 4). While in Zimbabwe the number increased owing to the reclassification of sub-IUs previously considered non-endemic or without data (Table 5).

The total target population for schistosomiasis in Ethiopia rose from 9.92 to 13.15 million (32.6%) and in Zimbabwe from 1.75 to 3.19 million (82.3%), representing substantial increases (Table 4). This increase reflects the expanded populations under the 2022 WHO guidelines, including pre-SAC, who were previously not treated, and adults, who were previously treated only where prevalence was ≥50%.

#### Scenario Three. Effect of a change in administrative unit of implementation

Scenario three uses the impact assessment data and the 2022 guidelines to evaluate the transitioning from IU to sub-IU implementation (Table 2). Overall, the number of sub-IUs requiring PC remain largely unchanged (4,414 to 4,544; 2.9%) and (1,714 to 1,723; 0.5%) for Ethiopia and Zimbabwe, respectively, while the total target population still increased across all age groups in both countries (Table 4). These increases arise because higher prevalence sub-IUs were classified at a lower prevalence when aggregated to IU-level.

#### Scenario Four. Effect of a change in WHO Guidelines

Scenario four assessed the effect of changing WHO guidelines, by comparing the 2011 and 2022 recommendations using sub-IU level impact assessment data (Table 2). Overall, there was a decrease in the number of sub-IU requiring PC in both countries by 24.8% in Ethiopia and 9.2% in Zimbabwe (Table 4). The 2011 guideline assigns biennial PC to all sub-IUs with 1–<10% prevalence, whereas the 2022 guideline replaces this with triennial or biennial PC or clinic-based test-and-treat where there was no previous PC, so fewer sub-IUs require PC.

Despite the reduction in geographic spread, the population requiring PC approximately doubled across both contexts, rising from 8.94 to 17.06 million (90.8%) in Ethiopia, and from 3.05 to 6.34 million (107.9%) in Zimbabwe. Adults, previously treated only under specific criteria, became eligible under 2022 guidelines, increasing by 10.78 million across both countries. Pre-SAC who were also previously ineligible, increased by 3.03 million in total. In both countries, a reduction in the number of SAC requiring PC fell alongside the geographic spread owing to the reduction in PC frequency from biennial to triennial (Tables 1, 5).

#### Scenario Five. Effect of a change in all decision-variables: epidemiological data, administrative unit plus WHO guidelines

Scenario Five changes all three decision-variables: data source, administrative unit and WHO guidance, the last including a lower prevalence threshold for treating adults and the addition of pre-SAC. This resulted in Ethiopia and Zimbabwe experiencing large increases in people requiring PC (Table 4).

In Ethiopia, sub-IUs meeting the treatment threshold fell by 42.5% (7,902 to 4,544), while the population requiring PC rose from 9.92 to 17.06 million (72.0%). Pre-SAC and adults requiring treatment expanded, but by comparison, SAC targets declined by 22.6% (9.01 to 6.97 million), consistent with reduced transmission in historically treated age groups.

In Zimbabwe, the number of sub-IUs requiring PC changed marginally (1,569 to 1,723; 9.8%). Pre-SAC and adults requiring treatment increased, while SAC requiring PC rose more modestly (1.66 to 2.55 million, 53.6%).

#### Scenario effects on PZQ requirements and treatment costs

Changes in PZQ tablet needs and delivery costs closely paralleled the population requiring PC trends (Table 4). Scenarios applying the 2022 guidelines, including pre-SAC and adult eligibility at ≥10% prevalence, produced the largest increases. Under Scenario Five, Ethiopia’s PZQ requirement rose by 78% and Zimbabwe’s by 307%, with delivery costs increasing proportionately: from US$1.98 to US$3.41 million in Ethiopia, and from US$0.77 to US$2.79 million in Zimbabwe.

## Discussion

Across two national programmes, we found that using a geostatistical approach for schistosomiasis impact assessments delivered sub-IU prevalence estimates with far lower survey effort and cost than a design-based approach, while increasing spatial coverage of data for programmatic decision-making. When those sub-IU data were paired with the 2022 WHO guidelines, the population eligible for PC, PZQ needs, and delivery costs rose substantially. These increased resources were driven mainly by the inclusion of adults and pre-SAC rather than by expansion of the number of areas treated. Our first and third hypotheses were supported. The second was not, moving decisions from the IU to the sub-IU changed the number of sub-IUs requiring PC by less than 3% in either country (Scenario Three); the reduction in geographic extent came instead from the change in guidance (Scenario Four). In brief, measurement efficiency through a geostatistical approach reduces survey inputs, and increases decision efficiency through sub-IU targeting, but eligibility expansion is the dominant driver of programme scale.

These findings are consistent with published evaluations of survey design [22,35]. Compared on the same simulated surfaces, model-based geostatistical analysis substantially outperforms the WHO guideline design for correctly classifying the endemicity class of an implementation unit, in three settings including Zimbabwe [22]. Design-based sampling scaled to the sub-IU - five schools of 50 children per sub-district - correctly classified a median of approximately 90% of sub-districts, but at the highest survey cost per correctly classified school among the designs evaluated [11]. Our study extends this work by carrying the consequences through to treatment requirements and delivery costs. Increasingly, studies demonstrate that moving to sub-IU targeting reveals meaningful heterogeneity and can reduce overtreatment [11–13,19]. Our results operationalise the WHO NTD Road Map’s call for sub-district, data-driven targeting that limits under- and overtreatment, ensuring programme equity and sustainability by more precisely identifying underserved populations [5]. Given the challenging global funding landscape [36,37], in which national NTD programmes have postponed or cancelled MDA, these savings are particularly compelling and may create opportunities for programmes to redirect resources toward additional programme activities including targeting more age-groups.

The expansion in programme scope we observe under the 2022 guideline reflects the broader objective of achieving elimination as a public health problem [5]. Evidence from modelling studies indicate community-wide treatment can accelerate reductions in prevalence and intensity compared with school-based strategies [38,39]. Reports of substantial untreated burden among adults in endemic settings reinforce the public-health rationale for expansion [40]. In this context, our results provide programme-level quantification from two countries, showing how policy and design choices translate directly into budgets and operational planning [41].

Several caveats merit consideration. The geostatistical approach relies on spatial interpolation; prediction uncertainty increases in data-sparse areas, and conservative exclusion rules may bias some estimates downward in inaccessible locations. Cost estimates reflect financial delivery costs and do not include all economic inputs (e.g., training, supervision, infrastructure, health system strengthening, opportunity costs, MoH overheads), thus underestimating the true costs. WorldPop population estimates are subject to uncertainty, particularly in remote areas or rapidly changing populations, which may affect estimates of the population requiring preventive chemotherapy and programme costs. Scenario analyses applied WHO algorithms consistently and did not simulate country-specific deviations, phased rollouts, or supply-chain constraints; thus, implementation realities may differ. These findings may be relevant to other schistosomiasis-endemic countries undertaking impact assessments and adopting the 2022 WHO guidelines as they transition towards elimination. How much the eligible population increases is likely to depend on the size of the already-treated population relative to the total population in areas meeting the treatment threshold: in Ethiopia, where a large school-based programme was already established, the eligible population rose by 72%, whereas in Zimbabwe, where a proportionally smaller population was being treated, it rose by 262%. However, the magnitude of these effects will vary according to local epidemiology, health system capacity, and programme delivery structures, and generalisability should be tested further before policy adoption. Finally, limited institutional capacity in geostatistics constrains scale-up, and complex or uncertain model outputs can be hard for non-specialists to interpret—delaying decisions—so building capacity, increasing modelling transparency, and engaging stakeholders are essential to improve uptake.

The findings are also conditional on the assumptions applied. Where the 2011 guidance specifies “maintain previous frequency” without a frequency, we applied annual PC; a less intensive interpretation would reduce the population attributed to the 2011 guidance and therefore increase, rather than decrease, the expansion we attribute to the 2022 guidance. The interpretations applied to the WHO algorithms follow WHO/ESPEN’s CDAT [34], although countries may deviate from these at verification, and the rule aggregating sub-IU classifications to IU level is a programmatic choice, so alternative thresholds would alter the IU-level scenarios though not Scenarios Four and Five. Delivery costs enter as a single cost per delivery, so a proportional error in that figure or in the population denominators shifts all scenario costs proportionally and leaves the comparisons between scenarios unchanged. The survey cost comparison is insensitive to unit costs because both survey types were costed from the same unit costs, so an error in any price affects both approaches and largely cancels; the exception is technical support, which is higher for the geostatistical approach and therefore narrows the reported difference (Supplementary Table S2).

These results identify two complementary aspects for improving programme efficiency and impact. First, measurement: a geostatistical approach provides precise, cost-effective data that supports decision-making and allows resources to shift from data collection to delivery without compromising accuracy. Second, implementation: moving treatment decisions from IU to sub-IU improves treatment efficiency by allowing more granular, evidence-based targeting that accounts for spatial heterogeneity.

In our scenarios, this refinement increased the population requiring PC, though less than the effect of the WHO guidance, by identifying persistent or emerging hotspots previously masked by IU-level aggregated prevalence. This approach is likely to be embedded in ESPEN microplanning tools which integrate historical epidemiological data, environmental suitability, and local knowledge [34,42]. Since the 2022 WHO guidance expands eligibility beyond SAC, national drug needs and delivery budgets may rise even if the number of areas treated remains stable or declines, particularly due to adult inclusion.. This study does not fully consider the additional health system capacity that may be required to support expanded treatment eligibility, including workforce, supply chains, monitoring systems, and community engagement. Therefore, future operational research evaluating the real-world implementation, feasibility, cost-effectiveness, and long-term sustainability of these recommendations in different endemic settings would further knowledge on efficiency gains for schistosomiasis treatment programmes.

## Conclusion

The geostatistical approach provides a cost-efficient route for generating the fine-scale evidence required for programme decision-making. Implementation of the 2022 WHO guideline at the sub-IU level substantially expands treatment eligibility, primarily because of changes in eligibility criteria rather than geographic expansion. Countries planning for elimination as a public health problem should pair efficient measurement with granular implementation and realistic budgeting for expanded eligibility to maximise health impact and avoid systematic under-treatment. Realising these gains depends on two conditions that not all programmes currently meet: access to epidemiological data of sufficient quality and spatial resolution to support sub-IU decisions, and the implementation capacity to deliver treatment at that resolution.

## Data Availability

All data produced in the present study are available upon reasonable request to the authors

## Authors’ contributions

Conceptualization: AC, FMF.

Methodology: AC, CF, ND, FMF.

Formal analysis: AC, CF, ND.

Investigation: FS, GTL, IP, NM.

Resources: FS, GTL, IP, NM.

Data curation: AC, ND.

Visualization: AC.

Funding acquisition: FMF.

Project administration: AC, FMF.

Supervision: FMF.

Writing – original draft: AC, CF, ND, FMF.

Writing – review & editing: AC, CF, FS, GTL, IP, ND, NM, FMF.

## Acknowledgements

We thank Alison Ower of the END Fund for her contribution and review of the original analysis and manuscript.

## Data Availability

The data used in this study come from the following sources.

**Zimbabwe**. Ward-level prevalence classifications from the national geostatistical impact assessment are publicly available in the supplementary materials of [24] (Supplementary Material 2, Table S7), with the underlying survey data and analysis code in Supplementary Materials 3–8. Baseline mapping data are published in [21].

**Ethiopia.** Kebele-level prevalence classifications from the national geostatistical impact assessment [23], and woreda-level baseline mapping data [20] obtained from the Joint Request for Selected Preventive Chemotherapy Medicines (JRSM) submitted by the Federal Ministry of Health, are owned by the Federal Ministry of Health of Ethiopia (FMoH) and the Ethiopian Public Health Institute (EPHI) and cannot be shared publicly by the authors. Permission to use these data for this study was granted by the FMoH and EPHI. Data were made available to the authors through their collaboration with the FMoH and EPHI; other researchers may apply for access through the Director, Infectious Diseases Research Directorate, Ethiopian Public Health Institute, Addis Ababa, Ethiopia.

**Population estimates.** Population estimates are publicly available from WorldPop (Constrained Individual Countries 2020, 100 m resolution; https://www.worldpop.org).

**Costs and outputs.** Survey costs by cost category are provided in S1 Table, and the cost per delivery used for each country is reported in the Methods. All scenario outputs are provided in Tables 3–5 and S2–S3 Tables.

## Supporting Information

Supplementary S1 Table: Differences in total survey costs (USD) by cost categories and survey type in Ethiopia and Zimbabwe to determine sub-implementation unit (kebele and ward, respectively) prevalence.

Supplementary S2 Table: One-way sensitivity analysis of the difference in total survey costs between design-based and geostatistical impact assessments in Ethiopia and Zimbabwe.

Supplementary S3 Table: Absolute and percentage change in the population requiring preventive chemotherapy for schistosomiasis, by target age group, in Ethiopia and Zimbabwe under five scenarios.

