## Supporting Information Tables S1-3 for "From district to community: expanded eligibility, not wider geography, drives schistosomiasis programme costs under revised WHO guidance in Ethiopia and Zimbabwe"

S1 Table. Differences in total survey costs (USD) by cost categories and survey type in Ethiopia and Zimbabwe to determine sub-implementation unit (kebele and ward, respectively) prevalence.

| Country | **Ethiopia** | | | **Zimbabwe** | | |
| --- | --- | --- | --- | --- | --- | --- |
| Survey type | *Design ($)* | *Geostatistical ($)* | *Difference (%)* | *Design ($)* | *Geostatistical ($)* | *Difference (%)* |
| Salary, per diems | 3,929,400 | 508,263 | -87.1% | 1,291,350 | 171,500 | -86.7% |
| Technical support | 123,563 | 519,526 | 320.5% | 66,815 | 162,095 | 142.6% |
| Communication | 1,500 | 1,134 | -24.4% | 1,200 | 1,900 | 58.3% |
| Training | 256,680 | 214,756 | -16.3% | 81,360 | 125,519 | 54.3% |
| Transport and fuel | 2,815,200 | 376,592 | -86.6% | 865,696 | 30,843 | -96.4% |
| Capital items | 1,073,550 | 23,846 | -97.8% | 192,147 | 3,840 | -98.0% |
| Survey materials | 2,761,329 | 250,201 | -90.9% | 72,132 | 204,026 | 182.9% |
| Sensitisation, social mobilisation | 131,820 | 44,197 | -66.5% | 17,291 | 4,800 | -72.2% |
| Results dissemination | 92,340 | 64,884 | -29.7% | 10,000 | 11,400 | 14.0% |
| Overhead | 671,123 | 263,411 | -60.8% | 805,377 | 221,936 | -72.4% |
| Total (w/o overhead) | 11,185,382 | 2,003,398 | -82.1% | 2,597,991 | 715,922 | -72.4% |
| Total (incl. overhead) | 11,856,505 | 2,266,809 | -80.9% | 3,403,368 | 937,858 | -72.4% |

Design: design-based impact assessments; Geostatistical: geostatistical impact assessments; USD: United States Dollar ($)

**S2 Table.** One-way sensitivity analysis of the difference in total survey costs between design-based and geostatistical impact assessments in Ethiopia and Zimbabwe.

| **Parameter varied** | **Ethiopia** | | **Zimbabwe** | |
| --- | --- | --- | --- | --- |
|  | **Difference (%)** | **Largest change (pp)** | **Difference (%)** | **Largest change (pp)** |
| **Baseline (Table S1)** | **-80.9** | **–** | **-72.4** | **–** |
| *Cost categories varied individually (±25%)* | | | | |
| Salary, per diems | -81.4 to -80.3 | 0.6 | -73.7 to -70.9 | 1.5 |
| Technical support | -81.9 to -79.8 | 1.0 | -73.5 to -71.4 | 1.1 |
| Communication | -80.9 | <0.1 | -72.4 | <0.1 |
| Training | -81.2 to -80.5 | 0.4 | -73.2 to -71.7 | 0.8 |
| Transport and fuel | -81.2 to -80.5 | 0.4 | -73.9 to -70.8 | 1.6 |
| Capital items | -81.3 to -80.5 | 0.4 | -72.8 to -72.1 | 0.4 |
| Survey materials | -81.4 to -80.3 | 0.6 | -73.8 to -71.1 | 1.4 |
| Sensitisation, social mobilisation | -80.9 to -80.8 | <0.1 | -72.4 | <0.1 |
| Results dissemination | -81.0 to -80.8 | 0.1 | -72.5 to -72.4 | 0.1 |
| Overhead | -81.2 to -80.6 | 0.3 | -72.4 | <0.1 |
| *Combined and alternative assumptions* | | | | |
| All cost categories varied simultaneously (±25%) | -80.9 | 0.0 | -72.4 | 0.0 |
| Technical support (±50%) | -83.0 to -78.8 | 2.1 | -74.6 to -70.4 | 2.1 |
| Wastage rate on capital items (0–20%; base 10%) | -81.0 to -80.7 | 0.1 | -72.6 to -72.3 | 0.1 |

Values are the percentage difference in total survey cost of the geostatistical impact assessment relative to the design-based impact assessment (negative indicates lower cost). Parameter difference (%) ranges give the values at the lower and upper bounds, and a single value is shown where these round to the same figure. pp: percentage points; largest change: maximum absolute deviation from the baseline difference; <0.1: non-zero change below 0.05 pp.

**S3 Table.** Absolute and percentage change in the population requiring preventive chemotherapy for schistosomiasis, by target age group, in Ethiopia and Zimbabwe under five scenarios.

| **Country** | **Target age group** | **Scenario one** | **Scenario two** | **Scenario three** | **Scenario four** | **Scenario five** |
| --- | --- | --- | --- | --- | --- | --- |
|  |  | **b − a** | **c − a** | **e − c** | **e − d** | **e − a** |
| *Ethiopia* | Pre-SAC (<5 yrs) | 0.00 (NA) | 1.73 (NA) | 0.68 (39.3%) | 2.41 (NA) | 2.41 (NA) |
|  | SAC (5–14 yrs) | −4.14 (−46.0%) | −3.10 (−34.4%) | 1.06 (17.9%) | −1.92 (−21.6%) | −2.04 (−22.6%) |
|  | Adults (≥15 yrs) | −0.92 (−100.0%) | 4.60 (500.5%) | 2.16 (39.1%) | 7.63 (15,260.0%) | 6.76 (734.8%) |
| *Zimbabwe* | Pre-SAC (<5 yrs) | 0.00 (NA) | 0.23 (NA) | 0.39 (169.6%) | 0.62 (NA) | 0.62 (NA) |
|  | SAC (5–14 yrs) | 0.63 (38.0%) | 0.14 (8.4%) | 0.75 (41.7%) | −0.48 (−15.8%) | 0.89 (53.6%) |
|  | Adults (≥15 yrs) | −0.09 (−100.0%) | 1.08 (1,200.0%) | 2.01 (172.9%) | 3.15 (15,750.0%) | 3.08 (3,422.2%) |

Populations are in millions. Values give the absolute change followed by the percentage change, where percentage change = (new − old)/old × 100. Scenario definitions and the corresponding populations for each scenario are given in Tables 2 and 4 of the main text. NA indicates that the comparator population was zero, so a percentage change cannot be calculated; this arises where an age group was not eligible for preventive chemotherapy under the comparator scenario. Very large percentage changes arise where the comparator population was small but non-zero, and should be interpreted alongside the absolute change. Pre-SAC: pre-school-aged children; SAC: school-aged children.
